# Quantitative assessment of pregnancy outcome following recurrent miscarriage clinic care: a prospective cohort study

**DOI:** 10.1101/2021.04.27.21255854

**Authors:** Rebecca C Shields, Omar Khan, Sarah N Lim Choi Keung, Amelia Hawkes, Aisling Barry, Adam J Devall, Stephen D Quinn, Stephen D Keay, Theodoros N Arvanitis, Debra Bick, Siobhan Quenby

## Abstract

**Objectives:** To measure pregnancy outcome following attendance at a recurrent miscarriage service and identify factors that influence outcome.

**Design:** Prospective, observational electronic cohort study.

**Setting:** Participants attending specialist recurrent miscarriage clinic, within a tertiary centre, with a history of two or more pregnancy losses. The clinic serves a diverse population (33% of residents belong in a minority ethnic group and over 33% in low-income households). Participant data were recorded on a bespoke study database, ‘Tommy’s Net’.

**Participants:** 777 women consented to participate. 639 (82%) women continued within the cohort, and 138 were lost to follow up. Mean age of active participants was 34 years for women and 37 years for partners, with a mean of 3.5 (1-19) previous pregnancy losses. Rates of obesity, BMI>30 (maternal: 23.8%, paternal: 22.4%), smoking (maternal:7.4%, paternal: 19.4%) and alcohol consumption (maternal: 50%, paternal: 79.2%) were high and 55% of participants were not taking folic acid.

**Outcome measures:** Biannual collection of pregnancy outcomes (ongoing pregnancy, live birth, still birth, pregnancy loss prior to 24 weeks), either through prompted self-reporting, or existing hospital systems.

**Results:** 639 (82%) women were followed up. 404 reported conception and 106 reported no pregnancy, at least 6 months following registration. Of those that conceived, 72.8% (294/404) had a viable pregnancy. Analysis identified a conception of rate of over 80% and viable pregnancy rate of 60% two years after attending the recurrent miscarriage clinic. 30% of couples had potentially modifiable risk factors for miscarriage.

**Conclusions:** Tommy’s Net provides a secure electronic repository on data for couples with recurrent pregnancy loss and associated outcomes. The study identified that subfertility, as well as repeated miscarriage, contributed to failure to achieve live birth. Study findings can enable comparison of clinic management strategies and inform the development of a personalized holistic care package.

**Funder:** Tommy’s Charity

**Sponsor:** University Hospitals Coventry and Warwickshire (UHCW) NHS Trust

**Trial Registration:** International Standard Randomized Controlled Trial Number (ISRCTN) Registry ISRCTN17732518; https://doi.org/10.1186/ISRCTN17732518

**Ethics:** Approval from West Midlands-South Birmingham Regional Ethics Committee IRAS No: 213740, 2225751 REC Ref: 17/WM/0050: 17/WM/208

**Strengths and Limitations of this study (related to the method):**

- The ‘Tommy’s Net’ e-repository and associated database contains baseline and prospective pregnancy outcome data from the largest known population of couples with recurrent miscarriage in the UK.
- Time to conception and viable pregnancy can be calculated from this data using time to event analysis.
- Obtaining follow up data is challenging but can be improved by using a variety of data collection methods.
- Follow up data is only requested biannually, therefore this is an inevitable lag in data collection.
- Limited use of the English language can be a barrier for participants completing the initial lengthy questionnaire.

**Key points:**

- 20% of this recurrent miscarriage population do not conceive and two years after first consultation 40% have not had a viable pregnancy. Early identification of this group could help facilitate early referral to fertility services or targeted research.
- Miscarriage is physically and psychologically challenging. Some couples may decide not to try to conceive again because of this. Ensuring appropriate psychological support is essential.
- Preconception care is inadequate. Over one third of couples attend their initial consultation with modifiable risk factors known to impact on miscarriage. Tackling these should be a priority.
- Having a BMI over 30 and being a smoker is more common within this cohort in women that do not conceive. Targeting of these risk factors may improve conception rate.

## Introduction

Miscarriage, the loss of a pregnancy prior to viability (24 weeks gestation) is common, with 15% of pregnancies ending in miscarriage^1^. Most miscarriages are sporadic and occur before 12 weeks of gestation^2^. Recurrent miscarriage (RM) is defined as two or three (or more) consecutive miscarriages^3,4^. It is estimated that 3% of women experience two consecutive miscarriages, and approximately 1% suffer three or more consecutive miscarriages^5,6^. In recurrent miscarriage, the incidence of euploidic foetal loss increases with each additional miscarriage, and the likelihood of a future successful pregnancy gradually decreases^7^. Recurrent miscarriage is a debilitating disorder, associated with considerable psychological morbidity^8^.

European and national miscarriage care guidelines recognise the importance of providing good physical care and psychological support^3,4^ however there are no standardised outcomes to assess care within clinics. A systematic review by MMJ van den Berg and colleagues (2018)^9^, evaluating features of care that couples valued within miscarriage services, found that information giving, including explaining potential causes of pregnancy loss and planning for future pregnancies were identified as areas for improvement.

Accurate information following attendance at a recurrent miscarriage clinic is important for couples’ counselling, stratifying care and directing research. Whilst data does exist around outcomes in a recurrent miscarriage setting^2,10,11^ it requires prospective update from clinics working under ESHRE guidance^3^, including all couples regardless of their outcome and not only those who conceived or who participate within a research trial.

The Tommy’s National Centre for Miscarriage Research brings together an interdisciplinary Translational Medicine research grouping jointly at the University of Warwick, University of Birmingham and Imperial College London. The Centre is dedicated to research across all aspects of miscarriage and early pregnancy complications including medical, basic scientific, social and ethical issues. A secure electronic data collection tool and e-repository (with associated database), Tommy’s Net, has been developed to facilitate recording of participant data, including follow up^12^.

## Objectives

Our objective was to quantify the long term cumulative live birth rate after first attendance at a recurrent miscarriage clinic. A cohort of couples was developed, with prospective data collection of the medical and obstetric histories of both partners, investigation results and pregnancy and neonatal outcomes. The tool for collecting data on this cohort is designed to be used in multiple clinics so that success rates between clinics can be benchmarked. This should also allow clinics to support and assess new care pathways, identify areas needing further research, develop outcome prediction modelling and investigate new tests in future clinical trials.

## Methods

The e-repository and associated database has been developed over several years by a team with representation from University Hospital Coventry and Warwickshire (UHCW) NHS Trust and University of Warwick, Imperial College and University of Birmingham. The cohort was initiated at UHCW, but designed so other clinics can join.

### Sponsorship, Ethics, Data management and Information Governance

Sponsorship (from primary hospital Trust), ethical permissions (IRAS No: 213740, 2225751 REC Ref: 17/WM/0050: 17/WM/208) and adherence to information technology governance standards was obtained. The study database complies with the regulatory requirements for Good Clinical Practice.

### Patient and public involvement

An established patient and public involvement (PPI) group from within the Tommy’s centre at UHCW was consulted during initial protocol development. Two further PPI sessions with 10 service users, each including 9 women and 1 partner, where consulted to ensure follow up methods where acceptable to participants and to optimise response rates.

### Setting

This cohort was established within a specialist recurrent miscarriage clinic in a tertiary referral centre (UHCW) within the UK. Miscarriage care followed European Society of Human Reproduction and Embryology (ESHRE) guidelines^3^.

### Eligibility

All couples with a history of two or more pregnancy losses (including biochemical loss^1^, miscarriage, molar pregnancy, ectopic pregnancy and stillbirth) were eligible.

### Recruitment

Couples are referred to the recurrent miscarriage clinic by their General Practitioner. Signposting prior to referral can occur from other hospital departments (e.g., Early Pregnancy Assessment Unit, Acute Gynaecology, Fertility unit) or charities (e.g., Tommy’s, The Miscarriage Association). Couples are then sent information about Tommy’s Net by post along with a baseline questionnaire. At their first clinic visit a member of the research team explains Tommy’s Net and asks them to consent to storage of their data.

### Data Collection

Both partners complete initial baseline questionnaires including demographic details, obstetric and medical history. Investigation results, blood pressure and body mass index (BMI) are recorded by clinic staff and entered into Tommy’s Net (see supplementary data).

The Tommy’s Net e-repository and database system, used for data collection and storage in the study, is based on the CURe framework^13^, a modular system for collecting research data in secondary care settings. The framework includes methods for the standardised, flexible capture and storage of data. The system is intended to link to the participating centre’s clinical information systems where possible to access relevant data already collected, such as laboratory test results. Tommy’s Net includes a database to organise data collected as part of the study and a web application for healthcare professionals to use for data entry, review and use in clinic. Data in Tommy’s Net can be exported for analysis. The development of Tommy’s Net has seen continuous improvements based on feedback from clinicians, researchers and patients. The design of the system is intended to promote interoperability with existing hospital systems to allow researchers to use information already collected, collect pregnancy outcomes to benchmark clinics and allow researchers to identify high risk groups of patients for future research.

### Statistical analysis

Statistical analysis was performed using IBM SPSS Statistics. Time to event analysis was performed using Kaplan-Meier curves, a non-parametric method for assessing the probability of an event occurring over time. Multi-variant analysis was conducted using age, BMI, cigarette smoking status, alcohol consumption and use of folic acid.

### Retention and Pregnancy Outcomes collection

A variety of methods were assessed to collect patient reported pregnancy outcomes after the first clinic visit. Initially women were encouraged to self-report outcomes by telephoning the clinic, or completing an outcome collection form sent by email. Automated invitations to complete this survey are sent via SMS every six months requesting information for follow up. This invitation consists of a single use link allowing the research team to trace the responses back to the patient identifiable baseline information.

Further outcome data is collected through viability scan visits, which can be accessed following initial review in the recurrent miscarriage service, and using existing hospital systems. Researchers used a maternity database, Evolution©, and a local intranet service to improve follow up and to validate participant reported information.

Using a variety of methods to collect outcomes improves follow up rate, however this does require researcher vigilance to avoid duplicate data entry. 17.8% of participants are still lost to follow up, therefore more work is needed in this area to encourage continuous engagement of participants.

### Improving baseline data

In the first three months of recruitment, a number of couples (n=83) consented to the study but did not complete the baseline questionnaire. This resulted in their data being marked as ‘inactive’ within the database (i.e., consented to the cohort study but not returned initial baseline questionnaires). On receipt of the baseline questionnaires, participants are ‘activated’ and followed up six monthly (n=10/83 to date). Our process has been updated so critical data items are collected by the clinician, from all couples who consent before leaving the initial clinic appointment. Participants are no longer registered within the database until they have completed the initial baseline questionnaire.

### Improving pregnancy outcome data collection

Initial pregnancy outcome data collection was poor with only 25% reporting their outcome, mainly due to technical difficulties in filling electronic versions of the forms for the participants. The response rate has gradually improved with development of a text message system. This was followed by other improvements such as a series of changes to the text message wording, by including partners in the messages, and changing the timing of the texts (with the majority sent in the afternoon or evening). Reminder messages are sent after 48 hours and after one week (if no responses from the initial text are received). Changes have been informed by patient and public involvement (PPI) groups, which were used to understand further why participants fail to respond to follow up SMS text message. Some explained that once they had had a baby, they were busy with their baby and forgot to reply. Conversely, repeated reporting of no pregnancy, or miscarriage was felt to be disheartening, or less important. We hope through education and careful wording of the questionnaire the response rate will continue to improve.

These approaches have contributed to an increase in response rate and combined with data from existing hospital systems, the response rate for pregnancy outcomes was 82.2%.

Data linkage with a general practice database was not deemed useful, because few miscarriages are recorded on the local general practice databases. Furthermore, there was a lack of standardisation in pregnancy data in primary care, though automated links with both primary and secondary care electronic health systems are still planned. The maternity services database may provide a fruitful source of pregnancy outcome data in the future.

## Results

### Analysis of cumulative live birth rate

Between May 2017 and January 2020, 777 women (and 480 partners) who attended the recurrent miscarriage clinic completed a baseline questionnaire and consented for their data to be included in the database. One hundred and thirty-eight (17.8%) participants were lost to follow up (no response to SMS, or information obtained for hospital databases), therefore 639 women are active within Tommy’s Net. One hundred and thirty-four of these women are within six months of consenting to the study and have not yet received a scheduled SMS. Five of these women have reported conceiving out with the SMS system with the data captured through early pregnancy scan clinics. Of the active women, their mean age was 34 years (see table I) and mean number of previous pregnancy losses was 3.5 (range 1-19). Demographic characteristics including age, ethnicity, alcohol intake, folic acid use and previous live birth were not statistically different between participants who conceived and those who did not (table I). Statistically more participants who did not conceive smoked and had a BMI over 30.

**Table I:** Comparison of demographics for all active participants, participants that did not conceive and those that were lost to follow up

|  | Continuing in cohort | No pregnancy | Lost to follow up | P value |
| --- | --- | --- | --- | --- |
| **Number** | 639 | 106 | 138 |  |
| **Age** | 33.7 | 34.03 | 33.7 | 0.092 |
| **Ethnicity** | White: 84% (436/519)<br>Mixed: 2.1% (11)<br>Asian: 8.9% (46)<br>Black: 3.3% (17)<br>Other: 1.7% (9)<br>Unknown (120) | White 85.5% (65/76)<br>Mixed: 2.6% (2)<br>Asian: 6.6% (5)<br>Black: 3.9% (3)<br>Other: 1.3% (1)<br>Unknown (30) | White 83.5% (101/121)<br>Mixed: 1.6% (2)<br>Asian: 9.8% (12)<br>Black: 2.4% (3)<br>Other: 2.4% (3)<br>Unknown (17) |  |
| **Average no. of previous live birth** | 0.6 | 0.15 | 0.23 | 0.36 |
| **Average no. of previous miscarriages** | 3.5 | 3.6 | 3.43 |  |
| **BMI over 30** | 23.8% (n=126/530) | 30% (n=26/87) | 18.2% (n=20/110) | 0.001 |
| **Smoking Y/N** | Yes:41 (7.4%) | Yes: 12 (13.5%) | Yes: 18 (14.9%) | 0.001 |
| **Number** |  |  |  | 0.000 |
| **Alcohol Y/N** | Yes: 278 (50%) | Yes: 51 (58%) | Yes: 75 (60%) | 0.083 |
| **Units** | 5.54 (0.5-30) | 5.03 (0.5-35) | 5.38 (0.5-30) | 0.000 |
| **Folic acid** | Yes: 292 (45.5%) | Yes: 35 (47.17%) | Yes: 57 (41.9%) | 0.000 |

### Pregnancy results

Four hundred and four of these women reported conceiving. One hundred and six (16.6%) women reported no pregnancy at least six months following registration, 31 (4%) of whom are no longer trying to conceive. Of those that conceived 72.8% (294/404) had a viable pregnancy (215 live births, 1 stillbirth, remainder currently <24 weeks at time of initial analysis). Analysis of data exported from the database in January 2020, revealed a conception of rate of 81% after two years within the cohort and viable pregnancy rate (pregnancy over 24 weeks or live birth at time of export) of 60% two years after attending the recurrent miscarriage clinic (fig 1). Age does impact on time to conception and time to viable pregnancy, with women of 25-34 years being more likely to have a viable pregnancy two years after initial review than other age groups.

**Figure 1:**
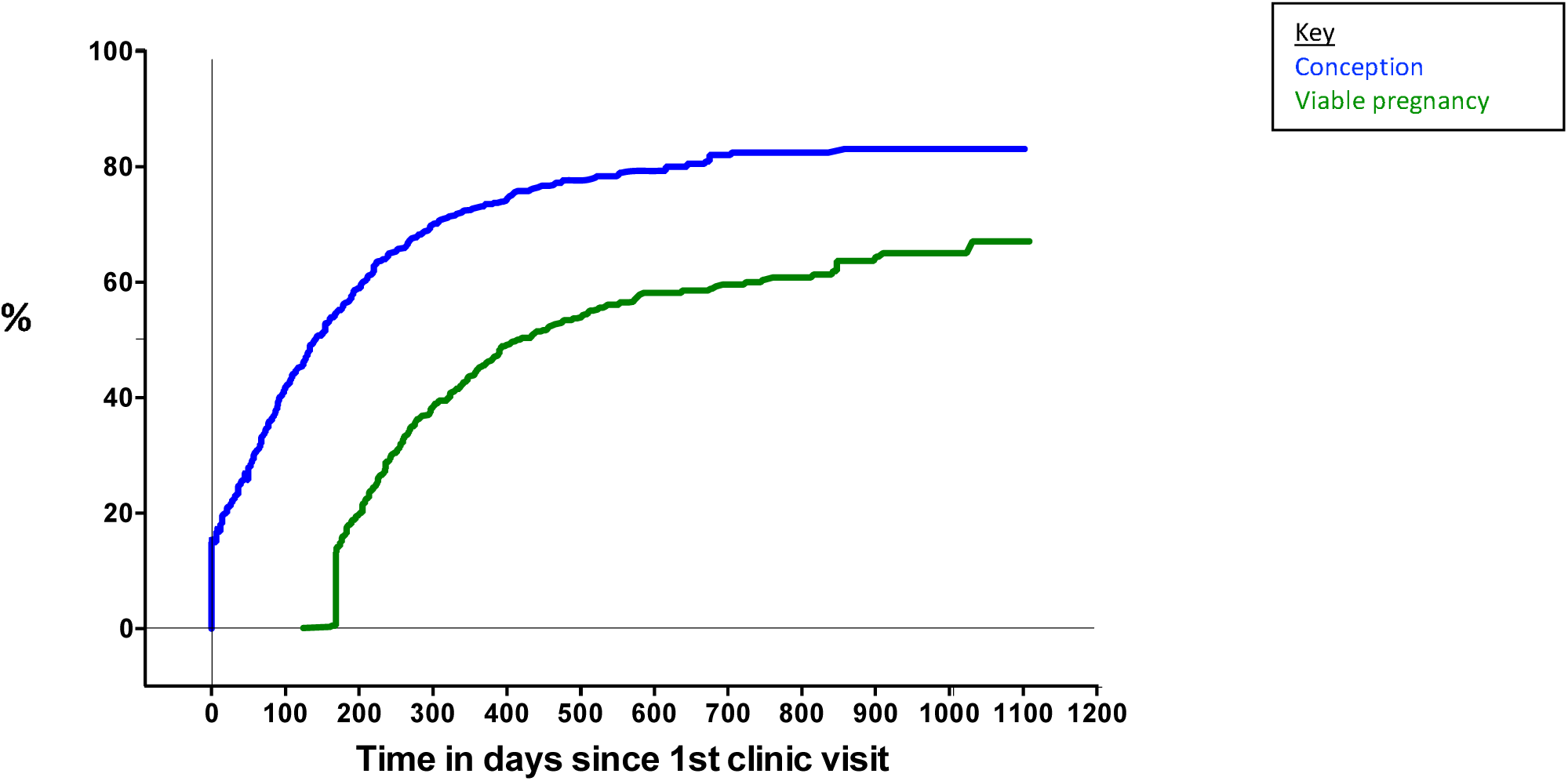
**Cumulative rate over time, from initial consultation to conception and viable pregnancy (>24 weeks gestation)**

The difference between couples who conceive and those who reach viable pregnancy starts at 30% (at 300 days conception rate is 70% and 40% reach over 24 weeks of gestation). This difference/gap gradually decreases and plateaus after 900 days to a difference of 19% (conception rate 82% with 63% reaching over 24 weeks gestation). The couples within this ‘gap’ represent those within our clinic who conceive but miscarry prior to viability despite current intervention and support. This gap is maintained within the 30-39years age group, but is less pronounced within those who conceive aged 25-29years (fig 2). Female BMI over 30 and female smoking status along with miscarriage history increases the time from initial consultation to conception and viable pregnancy within this patient group (fig 3-5).

**Figure 2:**
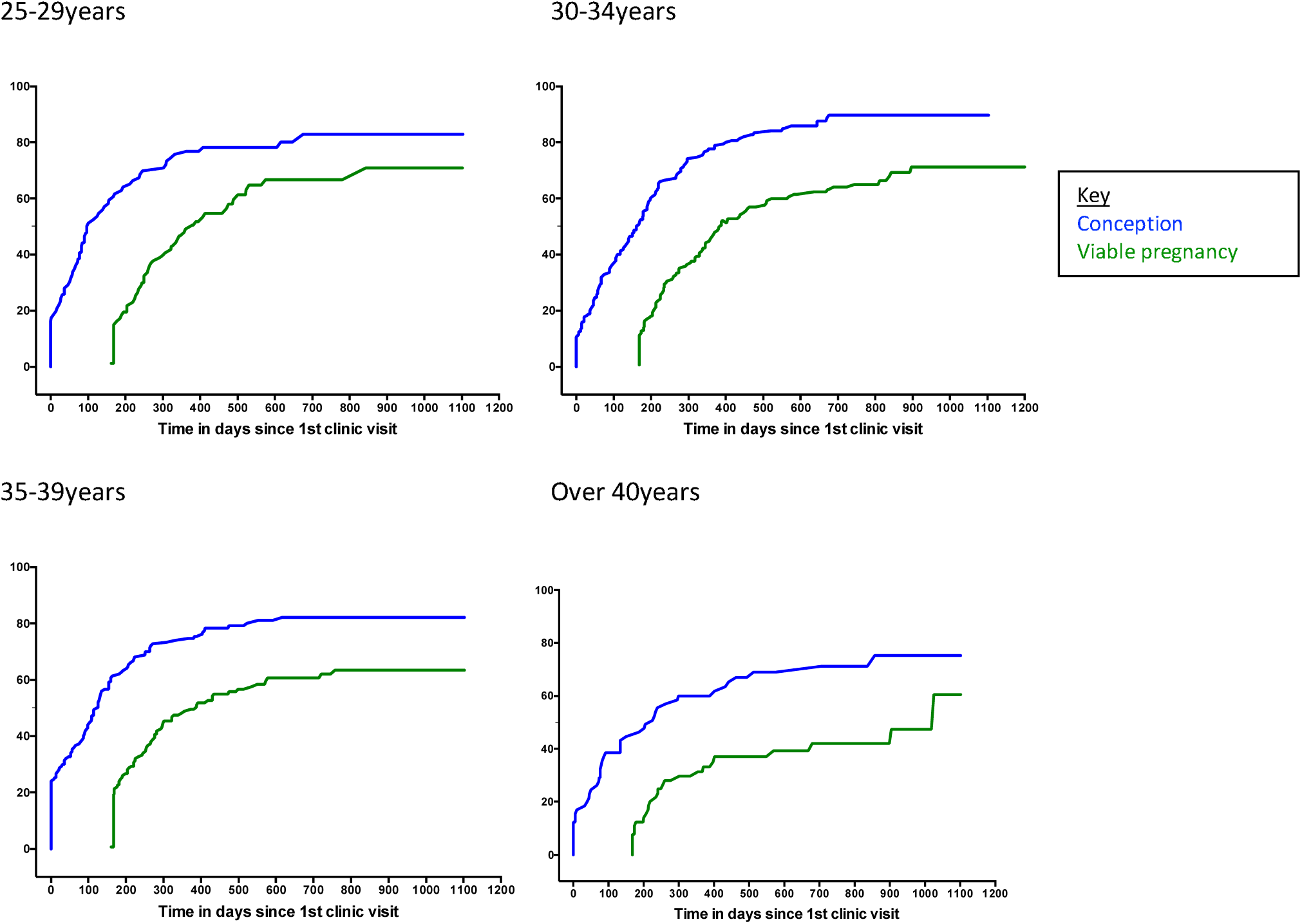
Comparing conception to >24weeks gestation by age.

**Figure 3:**
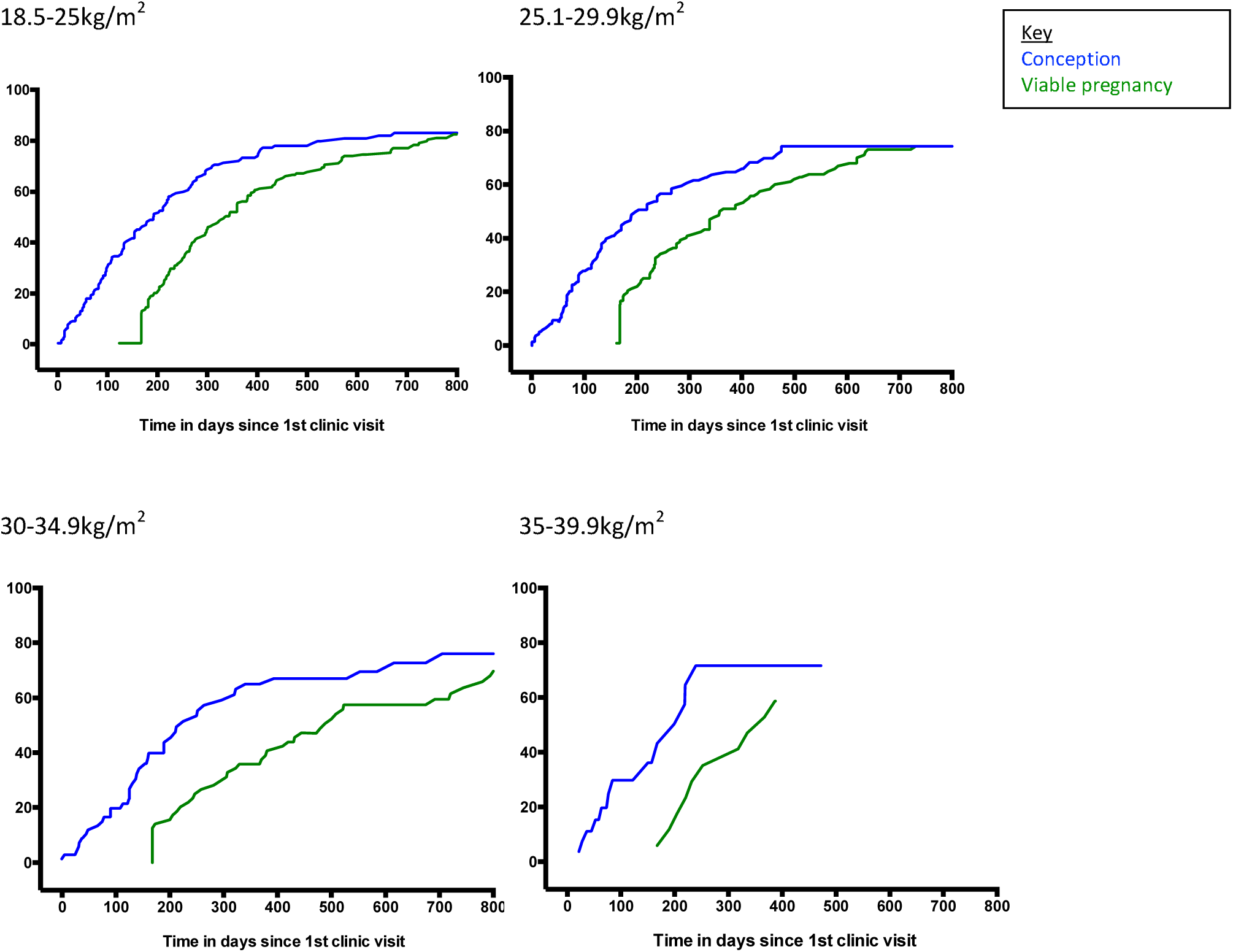
Time from initial consultation to conception/>24 weeks gestation by female BMI range.

**Figure 3a:**
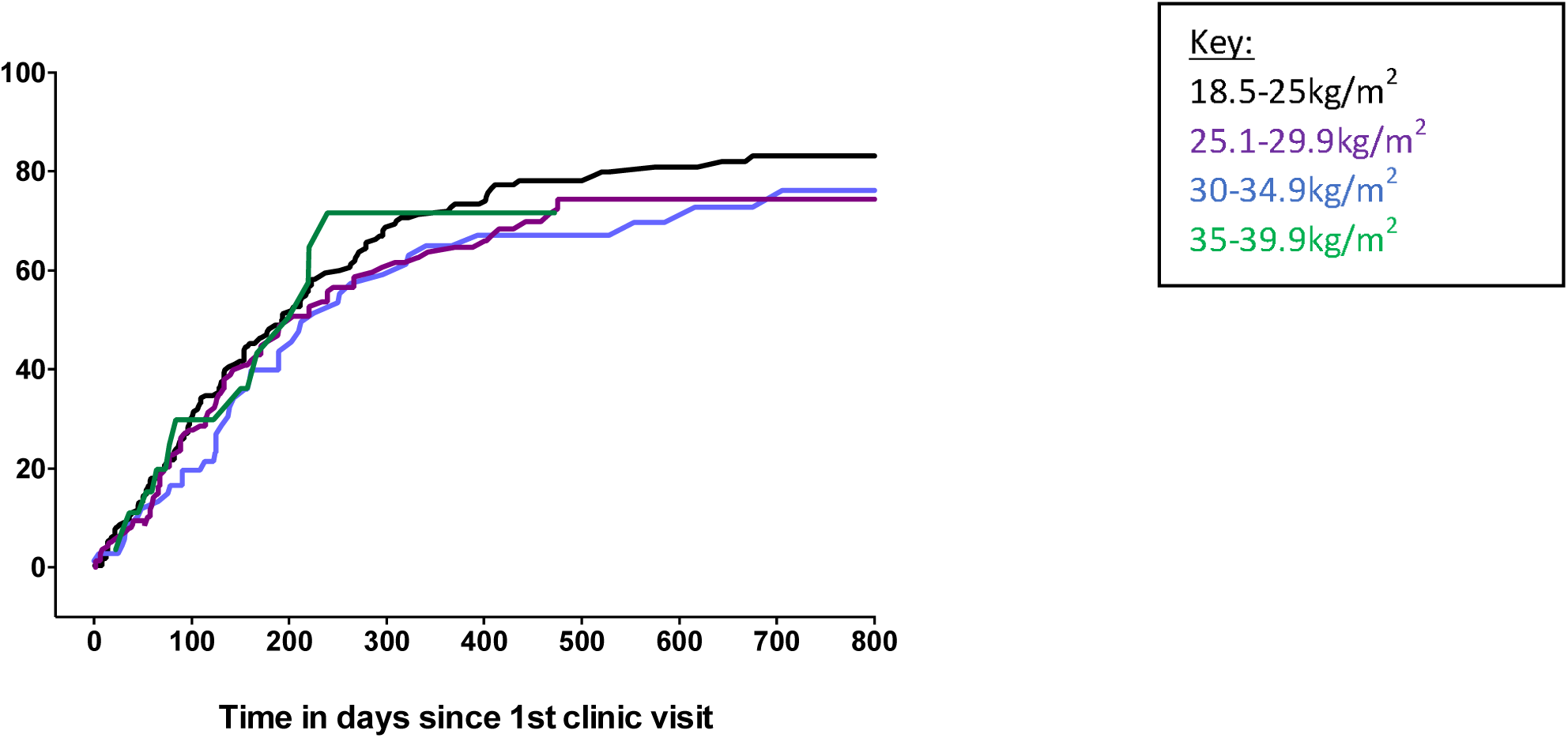
Time from initial consultation to conception by BMI.

**Figure 4:**
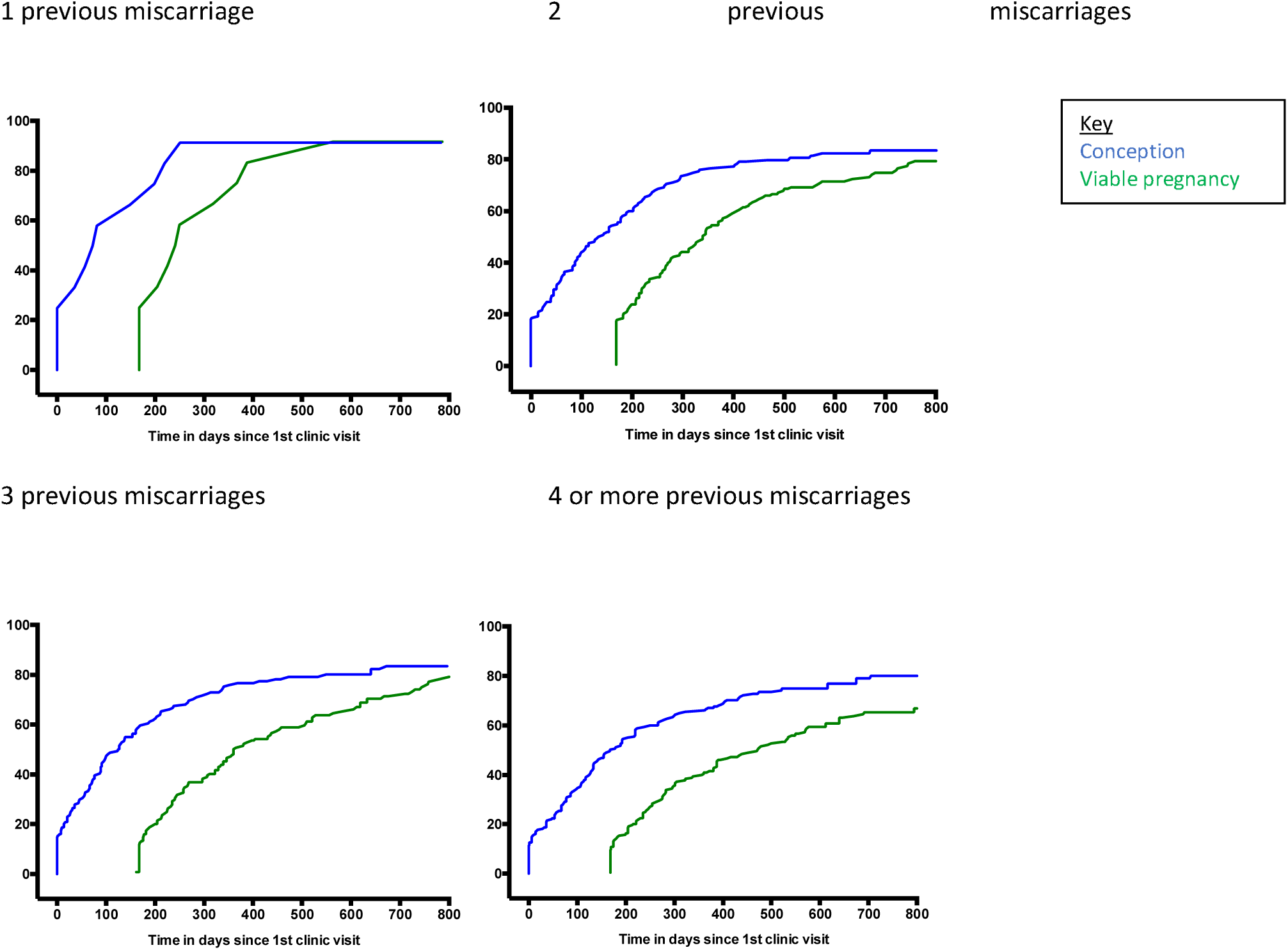
Time from initial consultation to conception/>24weeks gestation by miscarriage history.

**Figure 5:**
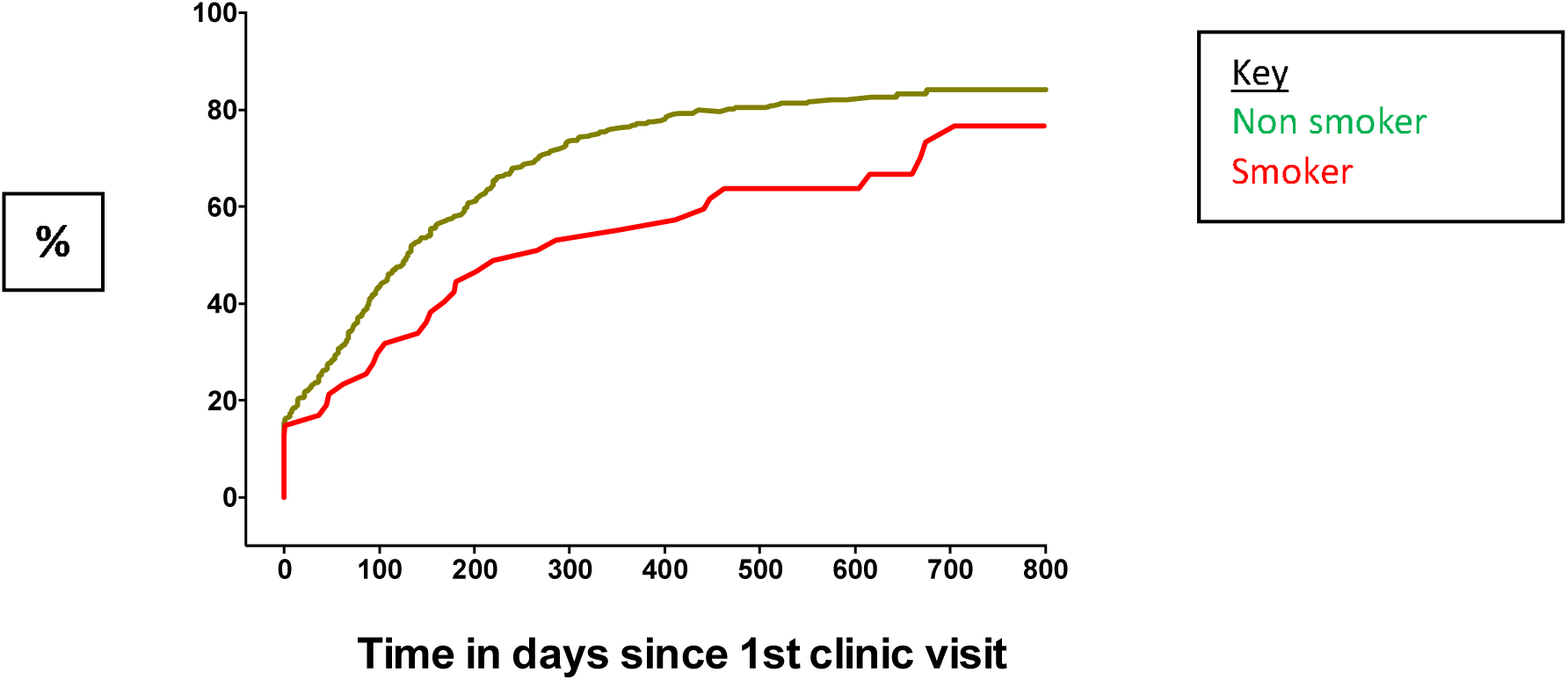
Time from initial consultation to conception by female smoking status.

A healthy BMI increases the chance of viable pregnancy, particularly when compared to a maternal BMI over 30kg/m2 (fig. 3). Having a BMI over 30 increases the time taken to viable pregnancy by 100-200 days. Within this population BMI does not appear to significantly change the time to conception (fig 3a), particularly within the first 300 days.

Couples who have had 4 or more miscarriages take longer to conceive, compared to couples who have has 3 or less miscarriages (fig. 4). There is a 17% gap within couples who have had 4 or more losses when comparing the rate of conception with viable pregnancy. This gap represents those that continue to miscarry and should be a population where research should be focused.

Smoking status impacts on time to conception. Females that smoke take longer to conceive with significantly more never conceiving.

## Discussion

### Database

We have developed an electronic method of obtaining outcomes from women following attendance at a recurrent miscarriage clinic. These outcomes can be used to assess recurrent miscarriage care and form a ‘benchmark’ to compare clinical services and interventions. The electronic cohort provides clinic outcome data in real time (on a dashboard, see supplementary data), and can be used for counselling couples as to both the chance of their next pregnancy succeeding and their cumulative time to live birth. This is novel, as data^2,10,11^ identified at literature review could not be generalised to the UK population. Lund and colleagues^10^ used a national, Danish registry to collect live birth data from attendees up to 5 years after their visit to a recurrent miscarriage clinic. Registry data were collected retrospectively and lacks information from couples who moved to other countries. Brigham^2^ analysed 716 couples over a 10-year period in their Liverpool clinic, with pregnancy outcome data on 325 patients with unexplained recurrent miscarriage. Data were only reported on those who conceived and had their pregnancy and birth care at the same hospital. These datasets are now over 20 years old. Kling and colleagues^11^ published more recent data based on a tertiary referral immunological centre within Germany. Seven hundred and nineteen couples were followed up for a median of 33.7 months, producing time to pregnancy and time to delivery over a five-year period. Whilst this is valuable data the study excluded couples who already had children within the partnership (25% within our clinic) and used immunotherapy in a proportion of couples which is not routinely used within the UK. It also asked for patient reported outcomes between nine months to four years after the event which could be prone to recall bias. This database will continue to collect and provide prospective outcomes of all those who attend this recurrent miscarriage clinic and, as use increases within the other sites it will allow comparison of outcomes with the aim of sharing good practice to improve patient care.

### Infertility

The time to conception curve within our RM population is similar to that in the general population^14^. Analysis to date has identified that within our cohort 16.6% (n=106) of couples fail to conceive within the follow up period. These patients are similar in age and ethnicity when compared to all within the active cohort. They do have a trend to a higher BMI, are statistically more likely to smoke.

Reasons why couples do not conceive are complex. Anecdotal evidence from the text message system and PPI groups shows some couples can feel unable to continue trying to conceive because of the potential risk of miscarriage. Recent research^15^ has documented an increased risk of post-traumatic stress disorder following pregnancy loss. We hypothesise that the psychological impact of miscarriage may stop couples from trying to conceive again. This is an important area in which to focus research and facilitate additional counselling and support.

Other couples may be unable to conceive despite actively trying. Identifying this subgroup of couples earlier could facilitate prompt referral to fertility services and hopefully increase their chance of conception and ultimately live birth. Within this population rate of conception decreases significantly 1 year after initial consultation (fig 1). 65% of couples conceive within 1 year of initial consultation, with only an additional 15% conceiving in the second year. In view of this decrease in pace of conception we suggest referral to fertility services should be considered within this population after 1 year.

Through-out the UK, access to NHS funded fertility treatment is dependent on maternal weight, smoking status, as well as age and parity. Addressing these factors early in the couple’s fertility journey may help to manage expectations prior to referral and reduce any delay in starting treatment. We recognise that weight particularly can be very sensitive issue and difficult to manage. Open and honest discussion, without blame, along with support and advice that joining group programmes for exercise and dietary modification can lead to more pregnancies than weight loss alone^16^ should be given. Referral to weight management services including dietetics and bariatric surgeons could be discussed if appropriate.

There may be a role for assessment of ovarian reserve within women with a BMI over 30, or who have previously waited over 12months to conceive. Having strong links, or an integrated multi-disciplinary preconception service may allow a more cohesive approach to these couples and increase their chance of having a viable pregnancy.

### Outcome Data

Comparing the ‘time to conception’ and ‘time to viable pregnancy’ curves illustrate the importance of assessing cumulative data. There is by definition a lag between conception and reaching 24 weeks pregnant, but following this the difference between the curves represents delay in live birth due to miscarriage. This gap decreases initially and may represent an impact from interventions and support within the recurrent miscarriage service. The importance of support to couples will be studied further during a planned qualitative study using semi-structured interviews of affected couples. After 900 days the gap between the curves is static and represents those whom despite conceiving have not yet had a child. This is a group which resources and research should be targeted to further understand reasons for miscarriage.

### Health Education

It is well documented that miscarriage risk increases with BMI over 30kg/m2 and smoking status^17-20^. Despite this 23.8% of women within the cohort have a BMI over 30kg/m2 and 7.4% smoke tobacco. Modifying these lifestyle factors through pre-conception counselling may reduce the chance of miscarriage and improve pregnancy outcome by reducing the incidence of, for example, gestational diabetes. Future research could be targeted at support in weight loss and smoking cessation.

### Limitations and strengths

The Tommy’s Net e-repository and associated database contains baseline and prospective pregnancy outcome data from the largest known population of couples with recurrent miscarriage in the UK. It allows calculation of ‘time to conception’ and ‘time to viable pregnancy’ using time to event analysis. This large dataset aims to facilitate future studies within a recurrent miscarriage population.

Obtaining follow up data is challenging. Using a variety of methods including self-reporting through the text message link and local hospital systems has improved our follow up rate.

Couples with limited English were unlikely to complete the lengthy questionnaire, which is currently only available in English. This means that this study may be missing high risk groups within our community

The introduction of the maternity services database could provide a valuable resource to enable improved follow up. Couples attend this RM clinic from all over the UK. Currently couples who deliver within our trust have at least two ways in which we can capture their outcome (SMS text message and hospital database with or without scan clinic information). These checks are not available to couples who have travelled some distance to attend and therefore may be under represented within the active participants group.

SMS text message requests for follow up are only sent every six months. This means that for the first six months that participants are within the study we do not expect to collect any outcome data. Some of these participants may go on to become ‘inactive’ and be removed from analysis.

## Conclusion

We have developed a user-friendly electronic database, storing comprehensive data, which can provide accurate time to conception and data on viable pregnancies to facilitate analysis into factors contributing to recurrent miscarriage. 16.6% of women within our clinic did not conceive and early referral to fertility services should be facilitated. Over 20% of women within the cohort have a BMI of over 30 and 7.4% smoke. Preconception counselling should be targeted at weight and smoking status with an aim of reducing miscarriage.

## Supporting information

Flowchart

## Data Availability

Original data may be obtained by emailing the corresponding author with a request.

## Authors roles

SQ had the initial concept. OK, SNLCK and TNA designed and developed Tommy’s net database and extracted initial data. RCS analysed the data and interpreted it along with SQ. RCS wrote the initial draft which was revised by SQ and DB, and reviewed by AH, OK, SNLCK, TNA, AB, AD, SDQ and SK. All commented on initial drafts and approved the final version.

## Acknowledgements

Thank you to all our participants, everyone in the Tommy’s Team at the Biomedical Research Unit, UHCW and Tommy’s for funding Tommy’s Net. Thank you also to all who participated in our PPI groups.

## Footnotes

1 Defined as no pregnancy identified on ultrasound scan

