## Supplementary figures and images for "Quantitative assessment of pregnancy outcome following recurrent miscarriage clinic care: a prospective cohort study"

### Flowchart

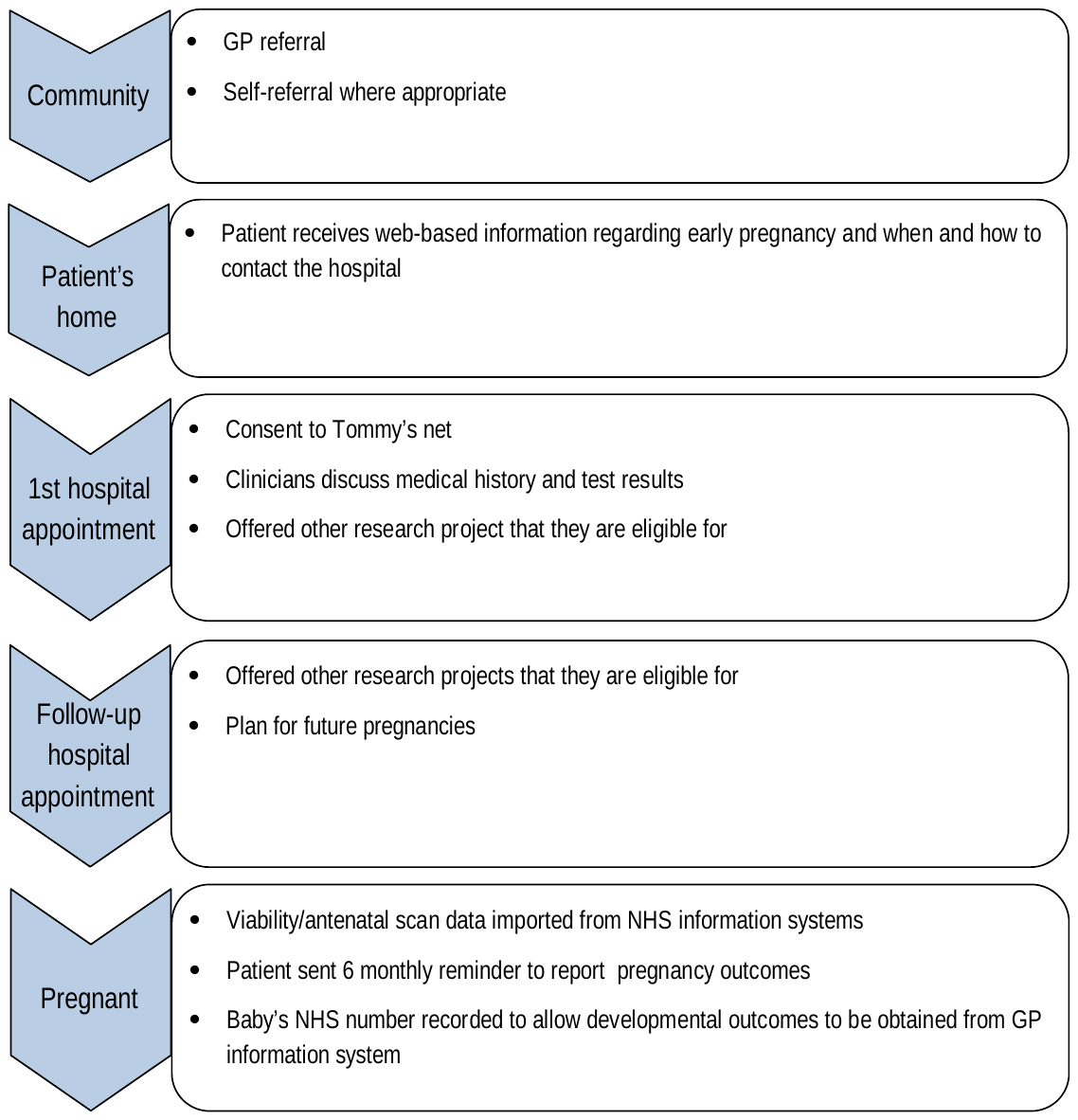
